# Unbiased human genomic characterization of polyglutamine disorder genes to guide biological understanding and therapeutic strategies

**DOI:** 10.1101/2025.02.17.25322412

**Authors:** Kevin Lucy Namuli, Britt I. Drögemöller, Galen E.B. Wright

**Affiliations:** Department of Pharmacology and Therapeutics, Rady Faculty of Health Sciences, University of Manitoba, Winnipeg, Canada; PrairieNeuro Research Centre, Kleysen Institute for Advanced Medicine, Health Sciences Centre and Rady Faculty of Health Sciences, University of Manitoba, Winnipeg, Canada; Department of Biochemistry and Medical Genetics, Rady Faculty of Health Sciences, University of Manitoba, Winnipeg, Canada

## Abstract

**Background:** Polyglutamine (polyQ) disorders, such as Huntington disease (HD) and several spinocerebellar ataxias, are severe neurological disorders caused by glutamine codon repeat expansions. These conditions lack effective treatments, with therapeutic research focused on pathogenic gene knockdown.

**Objectives:** We aimed to profile these genes using diverse human genomic data to inform therapeutic strategies by identifying new biology and assessing the potential on-target effects of knocking down these genes.

**Methods:** We conducted an unbiased phenome-wide study to identify human traits and diseases linked to polyQ disorder genes (Open Targets L2G>0.5). Network analyses explored shared trait associations and overlapping biological processes among these genes. Lastly, we assessed the theoretical druggability of polyQ disorder genes using recently identified features predictive of clinical trial success and compared them to repeat expansion (HD) modifier genes.

**Results:** We identified 215 human phenotype/polyQ disorder gene associations from 3,095 studies, indicating potential adverse effects from gene knockdown. Shared trait associations among genes suggested overlapping biological processes despite distinct functions. Drug target profile analysis revealed unfavorable risk profiles for polyQ disorder genes, particularly *ATN1*, *ATXN1*, *ATXN7*, and *HTT*, due to genomic features such as constraint, molecular interactions, and tissue specificity. PolyQ disorder genes also showed significantly more safety-related risks than HD genetic modifier genes (*P*=7.03×10^-3^).

**Conclusion:** Our analyses emphasize the pleiotropic nature of polyQ disorder genes, highlighting their potential risks as drug targets. These findings reinforce the importance of exploring alternative therapeutic strategies, such as targeting genetic modifier genes, as well as allele-selective approaches, to mitigate these challenges.

## Introduction

Polyglutamine (polyQ) disorders are a group of ten heritable neurodegenerative conditions characterized by repeat expansions of the glutamine codon in distinct genes.^1^ These include Huntington disease (HD), spinal and bulbar muscular atrophy (SBMA), dentatorubral-pallidoluysian atrophy (DRPLA), and several spinocerebellar ataxias (i.e., SCAs: 1-3, 6, 7 and 17).^1^ Collectively, they affect approximately 1-10 individuals per 100,000 people, contributing significantly to the increasing burden of neurological disorders globally.^2^

To date, some progress has been made in elucidating the pathogenic mechanisms of polyQ disorders. Unfortunately, these disorders remain without a cure, with current pharmacological treatments focused on alleviating the symptoms.^3,4^ While gene knockdown strategies, such as antisense oligonucleotides (ASOs), have been the focus of current research, and some of these ASOs have advanced to clinical trial stages, efficacy and safety issues remain substantial barriers to their clinical implementation.^3,5–7^ For example, the ASO tominersen demonstrated adverse events and failed to achieve efficacy. As tominersen suppresses the production of wild-type *HTT*,^8,9^ this disrupts normal gene function, highlighting the challenges associated with the therapeutic targeting of polyQ genes. This is particularly important given that all polyQ disorders present in an autosomal dominant manner, except for SBMA.

The role of adverse events in clinical trial failures underscores the importance of understanding the biological consequences of targeting specific genes when designing therapeutics.^10^ In line with this, Plenge *et al*.^11^ highlighted the significance of evaluating potential adverse clinical consequences *a priori* by identifying human phenotypes associated with genetic variation related to drug target genes.^11^ The increasing availability of extensive genome-wide association studies (GWAS) and related repositories, assisted by fine-mapping techniques that link genetic associations to suspected causal genes,^12,13^ allows for systematic and unbiased identification of gene-trait links. For example, we previously showed that common *HTT* genetic variation is associated with cognitive function in large human cohorts.^14,15^

Associations between polyQ disorder genes and human traits can also provide insights into the biological processes that these genes are involved in, aiding in the identification of additional potential therapeutic targets. Several studies have demonstrated that drugs with prior genetic evidence supporting the role that the targeted gene plays in the clinical phenotype being treated are more likely to succeed in clinical trials.^13,16–20^ Notably, a recent study also identified additional genetic factors associated with clinical trial outcomes,^19^ showing that genes displaying human genetic constraint, several interacting partners and broad gene expression were associated with clinical trial stoppage. These studies illustrate the importance of incorporating human genomic information into drug development pipelines.

The challenges highlighted above underscore the need to comprehensively evaluate polyQ disorder genes to identify potential on-target effects and investigate genomic features associated with therapeutic failures. Here, we aimed to characterize polyQ disorder genes using extensive human genomic data to inform therapeutic studies and disease biology. First, we analyzed and prioritized human trait and disease associations with common genetic variation in polyQ disorder genes using large repositories. We unbiasedly profiled important human genomic information through this approach to identify potential on-target effects. We also uncovered unique and shared biological features by examining phenotypes associated with these genes outside of the polyQ disorders themselves. Finally, we evaluated the theoretical druggability of the polyQ disorder genes, ranking them according to features related to clinical trial success and comparing them to the repeat expansion modifier genes that influence disease onset. Our comprehensive analysis enhances our understanding of the underlying biology of polyQ disorders and offers potential insights into the consequences of targeting the associated genes for polyQ disorder therapeutics. The findings from our study have broad relevance to therapeutic strategies for other repeat expansion disorders, of which more than 50 have been identified.

## Material and Methods

### Ethical approval and gene selection

Ethical approval was obtained from the University of Manitoba Bannatyne Research Ethics Board (H2022:354 HS25743). We profiled all known polyQ disorder genes (*n*=10, i.e., *AR*, *ATN1*, *ATXN1*, *ATXN2*, *ATXN3*, *ATXN7*, *CACNA1A*, *HTT*, *TBP* and *THAP11*). For GWAS-based analyses, we excluded *AR* because it is located on the X chromosome, and most current GWAS do not include this information.

### Identification of human traits and diseases associated with polyQ disorder genes

To identify human traits and diseases associated with the polyQ disorder genes beyond their primary disorders, an unbiased, comprehensive phenome-wide study was conducted by analyzing data from the Open Targets Genetics database (OTG) (version 8).^12^ This repository contains approximately 132,893 curated GWAS association signals from various studies and genomic biobanks (e.g., FinnGen and UK Biobank), including 8,317 human GWAS summary statistics (extracted January 2024). To curate OTG data, we used the related application programming interface, implemented using GraphQL v2.0^12^ and executed in R 4.3.3.

We accessed polyQ disorder gene data using Ensembl identifiers.^21^ We identified human traits where significant associations could be attributed to polyQ disorder genes using the locus-to-gene (L2G) fine-mapping tool.^22^ L2G is designed to refine genome-wide significant GWAS signals (*P*<5×10^-8^), identifying likely causal genes attributed to these signals.^12^ This tool uses a scoring metric that quantifies the strength of evidence for gene-trait associations, ranging from 0 to 1.^12^ Higher scores signify increased support for a particular gene-trait association. Traits overlapping with the polyQ disorder genes with L2G>0.5 were kept for further analysis as recommended by the developers of this tool.^22^

Finally, to examine other severe Mendelian disorders and other phenotypes associated with polyQ disorder genes outside of the canonical polyQ disorder, we profiled the Online Mendelian Inheritance in Man (OMIM) database.^23^ We analyzed the OMIM genemap2 dataset (accessed June 2022) containing 5,779 phenotype entries.

### Assessing shared traits among polyQ disorder genes

To investigate whether polyQ disorder genes are linked to shared traits and the degree of this overlap, we performed a network analysis using *ggraph* (version 2.1.0) and *arcdiagram* (version 0.1.12) in R. All gene-trait associations with an L2G>0.5 were included in the network analysis. In the resulting network, each node represented a polyQ disorder gene, and each edge (link) represented a bidirectional relationship shared between two genes. This network analysis aimed to identify networks of genes linked to particular traits, thereby providing insights into underlying mechanisms and shared biology.

### Prioritization of informative L2G-significant human trait associations for polyQ disorder genes

To prioritize individual traits and disease signals for the polyQ disorder genes, count-based traits (e.g., blood cell types) were removed. This was performed to minimize non-specific signals, as such traits are associated with numerous generic associations across the genome. Further, we restricted our analyses to include only gene-trait pairs derived from peer-reviewed PubMed studies to ensure the robustness of study findings.

### Variant annotation and unique signal identification

We annotated the predicted functional effect of index variants related to GWAS trait signals to understand their potential mechanistic contribution to the observed gene-trait associations. Ensembl Variant Effect Predictor was used to identify the *most severe* consequence flag related to each polyQ disorder gene. SpliceAI was used to predict whether the variants alter splicing.^24^ Genotype-Tissue Expression (GTEx) data were assessed to determine whether variants were linked to gene expression changes of the related polyQ disorder genes. Further, we identified independent signals at each gene by considering linkage disequilibrium (LD) between index variants. Independent signals, defined as haplotypes, were determined by pruning index variants with *r*^2^>0.5 using the 1,000 Genomes European super-population with LDLink SNPClip.^25^ Each haplotype was described as a set of variants in LD (i.e., *r*^2^>0.5) with each other. A single tag variant was considered a proxy for all the variants in high LD and was used to capture these haplotypes. The Finnish population was excluded from this analysis to minimize potential confounding factors from extended haplotype blocks.^26^ Resulting haplotypes were assigned to prioritized polyQ disorder gene-trait pairs to determine unique signals at each locus.

### Assessment of the theoretical druggability and risk profiles of the polyQ disorder genes

To thoroughly evaluate polyQ disorder genes for theoretical druggability and drug target profiles related to the risk of clinical trial failure, we assessed recently identified features associated with clinical trial success.^19^ Genes were categorized as favorable/unfavorable under each category based on previously determined thresholds.^19^ The following features were included:

(i) <u>Genomic constraint</u>: We examined genes’ tolerance to loss-of-function mutations using sequencing data from the Genome Aggregation (gnomAD) Database, which contains 125,748 exomes and 15,708 genomes (gnomAD v2.1.1; accessed July 2022).^27^ We used the probability of loss-of-function intolerance (pLI) metric, which ranges from 0 to 1, with more constrained genes having higher scores. Genes with a high degree of constraint were categorized as unfavourable, indicated by a rank greater than 0.9 (corresponding to *“extremely loss of function intolerant”* genes).^28^
(ii) <u>Tissue expression</u>: We accessed RNA tissue specificity data from the Human Protein Atlas (HPA) database (accessed February 2023),^29^ which uses data from the HPA and GTEx to classify genes into five categories (i.e., *“tissue enriched”*, *“group enriched”*, *“tissue enhanced”*, *“low tissue specificity”* and *“not detected”*) based on fold change in mRNA expression across 37 organ systems.^30^ Genes whose RNA tissue specificity was classified as *“low tissue specificity”* were ranked unfavorable as they are broadly expressed.
(iii) <u>Molecular interactions (MI)</u>: We used the Open Targets Platform to access data from the IntAct database^31^ related to MI evidence for the genes (accessed October 2024). We used the MI score, which ranges from 0 to 1 (scores closer to 1 are higher confidence) and is built using diverse data (i.e., publications, experimental detection methods, and interaction types).^32^ Genes with more than ten interacting partners with MI scores exceeding 0.42 (considered high confidence^19^) were unfavorable.
(iv) <u>Druggability evidence</u>: We assessed the Drug Gene Interaction Database (accessed July 2024)^33^ to identify any known or potentially druggable genes among the polyQ disorder genes. We assigned a favorable “*druggable any*” metric to polyQ disorder genes that had any known or predicted evidence of interaction with drugs from this database.

Finally, we compared the number of unfavorable genomic features associated with increased safety-related risk (i.e., constraint, expression and interactions) to these same metrics reported in our recent analysis of HD modifier genes^34^ using the Wilcoxon test accompanied by the calculation of the effect size (*r*) to quantify the magnitude of the observed difference. This analysis aimed to assess whether targeting one of these gene sets presents a theoretically greater safety risk than the other. *P*<0.05 was considered significant in these analyses.

### Data sharing

The data used in the analyses described in this study are publicly accessible to researchers through the consortia that generated them. Analysis scripts for data accession, analysis and visualization are accessible at https://github.com/Wright-Lab-Neurogenomics-Research/polyQ_genes_characterization_analyses

## Results

### Profiling polyQ disorder genes for associations with common human traits and diseases

We identified 3,095 association signals (1,467 unique traits) surrounding polyQ disorder genes from the OTG database. These traits were distributed across 20 categories based on health conditions or biological processes, with the broad *measurement* category containing the most associations (*n*=2,521). Applying a filtering threshold of L2G>0.5 retained seven polyQ disorder genes (i.e., *ATXN1*, *ATXN2*, *ATXN3*, *ATXN7*, *CACNA1A*, *HTT* and *TBP*) and identified 215 gene-trait pairs (149 unique traits), signifying associations with strong evidence for polyQ disorder gene involvement (**Figure 1**). *ATXN1* had the highest number of L2G>0.5 significant traits reported (*n*=57), predominantly related to hematological (measurement) phenotypes. *ATXN3* and *TBP* were exclusively linked to hematological phenotypes, while *ATXN7* was associated solely with disease-related phenotypes. Network analysis of the 215 polyQ disorder gene-trait pairs with L2G>0.5 revealed that five polyQ disorder genes shared phenotypic traits. *TBP* and *CACNA1A* did not share traits with any polyQ disorder genes. *ATXN1* had the greatest traits shared with the other polyQ disorder genes. *HTT* and *ATXN1* displayed the most shared traits, including educational attainment and cognitive ability, in addition to neutrophil and white blood cell counts.

**Figure 1.**
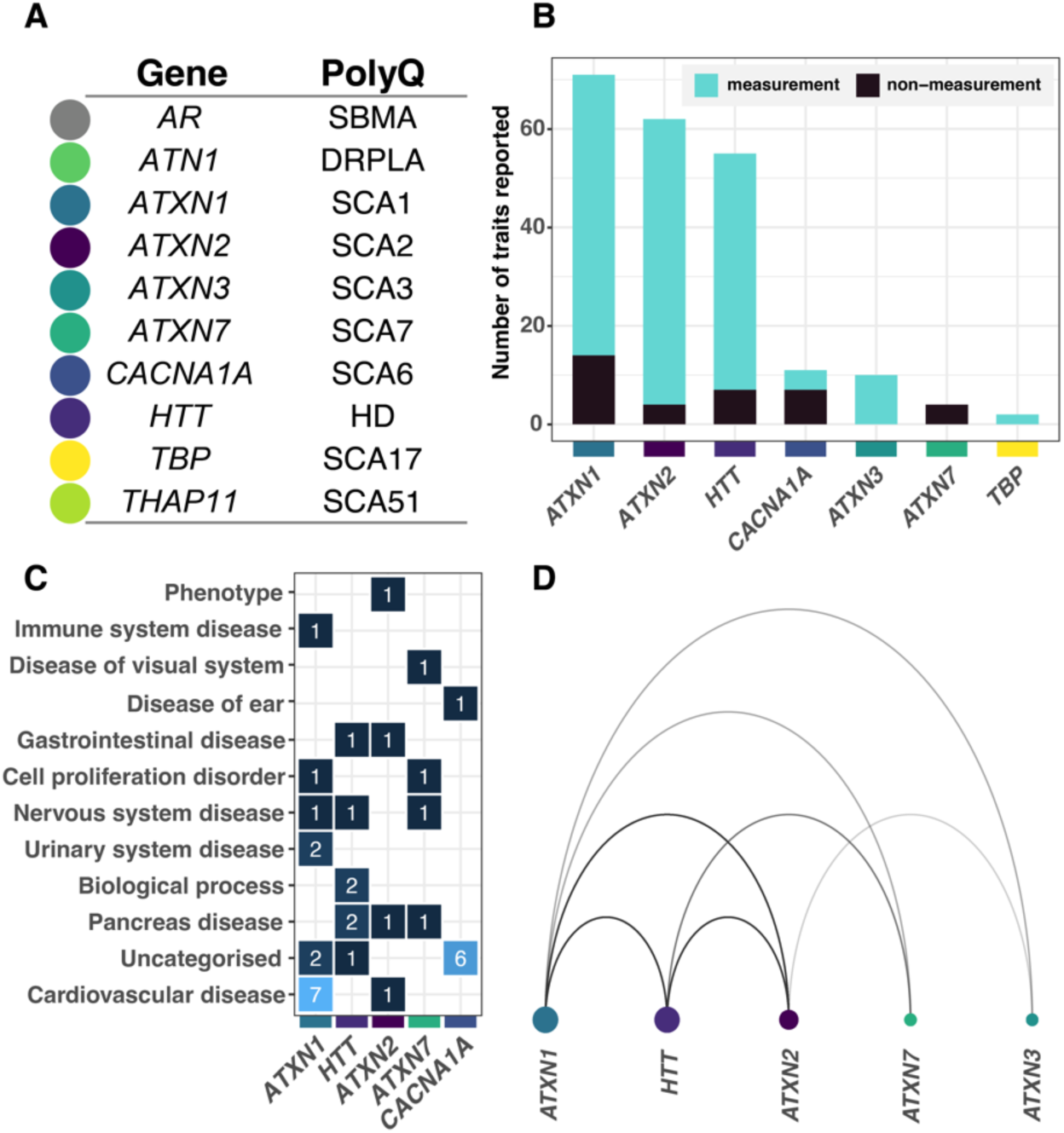
PolyQ disorder genes are pleiotropic and associated with several human traits and diseases in diverse phenotypic categories. **(A)** polyQ-associated genes and their corresponding polyQ disorders. Colors represent different genes used across panels and figures. **(B)** Human traits and disorders linked to polyQ disorder genes (i.e., L2G>0.5) obtained from an analysis of genome wide association studies in the Open Targets Genetics Database. The highest number of gene-trait pairs were identified for *ATXN1*, *ATXN2*, and *HTT* and most traits were classified as measurements. **(C)** Heatmap of non-measurement-related traits (L2G>0.5) for each polyQ disorder gene, depicting several associations for disease including those related to cardiovascular system (e.g., atrial fibrillation), pancreas (e.g., type 2 and type 2 diabetes) and nervous system. **(D)** Network graph illustrating shared polyQ disorder gene-trait associations. Larger circles represent the number of traits associated with a particular gene, while thicker arcs indicate more shared trait associations between genes. Notable shared traits include *HTT* and *ATXN1* with educational attainment and cognitive ability; *ATXN1* and *ATXN7* with breast cancer; and *ATXN1*, *ATXN2* and *ATXN3* with HDL cholesterol levels. This network highlights the pleiotropic effects of polyQ disorder genes across neurological, metabolic, and oncological traits. **Abbreviations:** L2G; locus-to-gene; *DRPLA*, Dentatorubral-Pallidoluysian Atrophy; *HD*, Huntington Disease; *SBMA*, Spinal and Bulbar Muscular Atrophy; *SCA*, Spinocerebellar Ataxia.

We applied further prioritization steps to obtain high confidence and informative traits linked to polyQ disorder genes, including removing count-based terms and studies not found in PubMed. These analyses retained 46 significant traits (L2G>0.5) and five polyQ disorder genes: *HTT, ATXN1, ATXN2, ATXN7*, and *CACNA1A* (**Figure 2**). Furthermore, filtering out duplicate traits for each gene retained 37 unique gene-trait associations. These informative trait associations represent diverse disease phenotypes, including neuropsychiatric traits (e.g., *HTT*-depression, *CACNA1A*-positive affect, *ATXN7*-schizophrenia), metabolic disorders (e.g., *ATXN2*-type 1 diabetes, *HTT/ATXN7-*type 2 diabetes) and autoimmune disorders (e.g., *ATXN1*-multiple sclerosis, *ATXN1*-systemic lupus erythematosus). Additionally, they include several non-neurological traits or diseases, such as *ATXN1*-atrial fibrillation, *ATXN1*-/*ATXN7*-breast cancer, and *ATXN7*-cataracts. PolyQ disorder genes, such as *ATXN1* and *HTT,* were associated with non-pathogenic neurogenetic traits, including educational attainment, cognitive ability, and aging. This collective observation highlights the pleiotropic features related to these genes outside of polyQ pathobiology.

**Figure 2.**
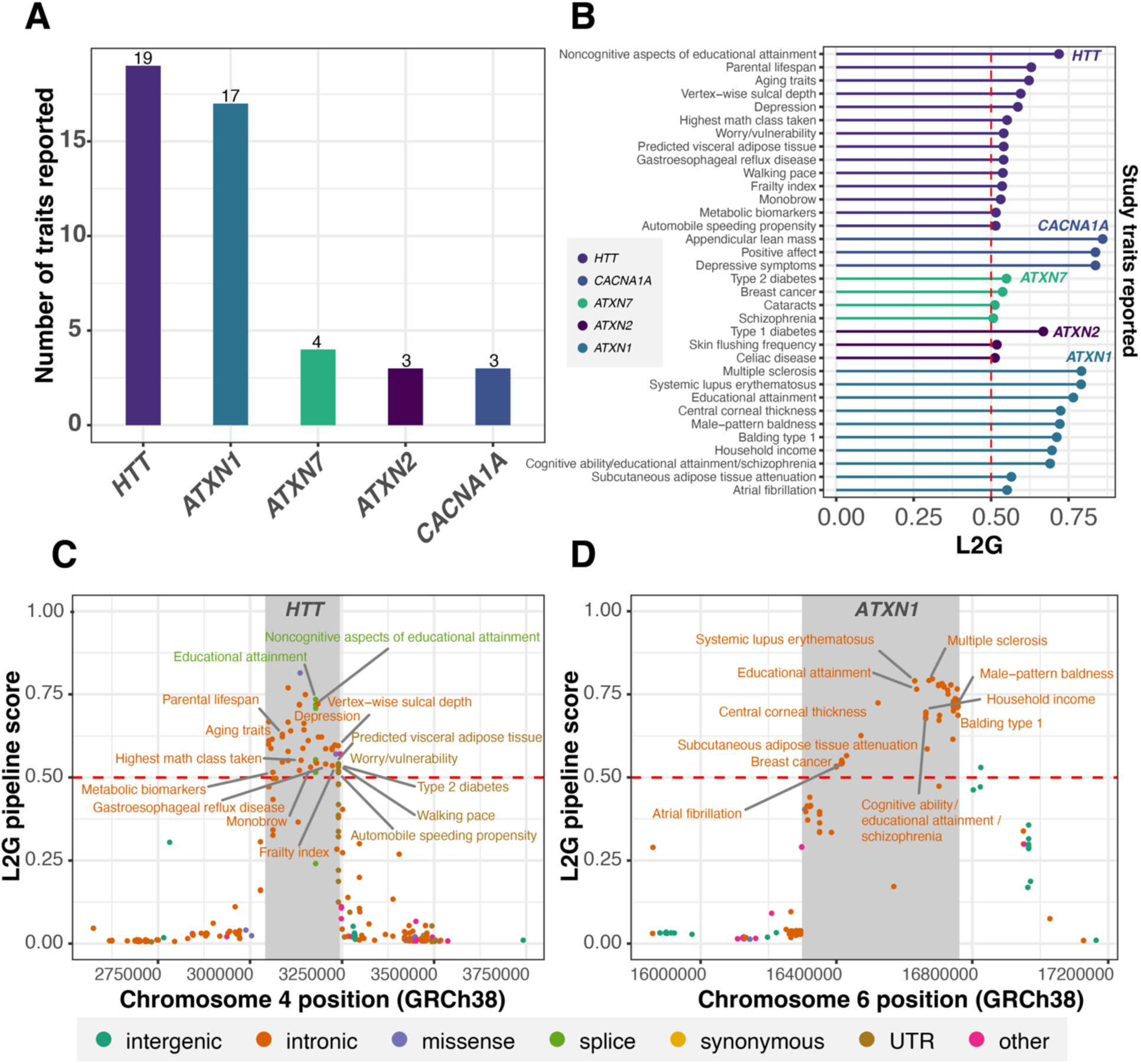
High-confidence and informative human gene-trait associations linked to the polyQ disorder genes. **(A)** A total of 46 traits linked to five polyQ disorder genes remained after applying a filtering threshold of L2G>0.5, while excluding count-based terms (e.g., blood cell counts) and studies lacking PubMed IDs. (**B)** L2G scores for individual high confidence trait associations (duplicate traits removed). PolyQ disorder genes exhibit associations with both pathogenic and non-pathogenic characteristics, across neurological, metabolic, and immune-related traits. **(C)** *HTT* and **(D)** *ATXN1* displayed the highest number of gene-trait pairs and are linked to neurological disorders and neurobiological phenotypes. In these panels, high confidence gene-trait associations are annotated and plotted by variant position and L2G score. Color indicates predicted variant consequence (*most severe*) of index variant for each trait association. Count based traits were plotted but not annotated. The grey-shaded segment represents polyQ disorder gene locus (GRCh38-genome build coding sequence start-stop position). **Abbreviations:** L2G; locus-to-gene, UTR; Untranslated region, GRCh38; Genome Reference Consortium Human Reference 38.

### Investigation and annotation of the variants linked to unique polyQ disorder gene-common trait associations

The index variants linked with the high-confidence unique traits associated with the polyQ disorder genes fall into three categories: intronic (*n*=26), splice (*n*=1), and variants located in untranslated regions (*n*=1) (**Figure 2**). Notably, all traits associated with the splice and untranslated region variants were annotated to *HTT*. In contrast, traits related to the remaining polyQ disorder genes (i.e., *CACNA1A, ATXN1, ATXN2* and *ATXN7*) were exclusively associated with intronic variants. The *HTT* splice variant showed associations with educational attainment and its noncognitive aspects. *ATXN7* was associated with several pathogenic traits, including type 2 diabetes, schizophrenia, breast cancer and cataracts, each associated with a unique variant (total *n=*4).

The 37 unique gene-trait associations arose from 30 distinct signals. A variant located in the untranslated region in *HTT* was associated with the highest number of distinct traits (*n=*5), including type 2 diabetes and walking pace (**Table 1**). The remaining unique variants resulted in 21 independent signals (i.e., *ATXN1, n*=8; *HTT, n*=6; *ATXN7, n*=3; *ATXN2, n*=2; *CACNA1A, n*=2) referred to as haplotypes. The *HTT* haplotype two (index variant: intronic rs61348208), which we have previously reported,^15^ had the largest number of traits (*n*=6) associated with it, including both pathogenic (e.g., depression) and non-pathogenic traits (e.g., aging traits and parental lifespan). *ATXN1* exhibited the highest number of independent signals (*n*=8 haplotypes), with *ATXN1* haplotype three (index variant: intronic rs909788) (*n*=5) demonstrating the most trait associations. However, these were all linked to highly correlated cognitive and socio-economic traits.

**Table 1.** High-confidence GWAS associations linked to polyQ disorder genes are associated with pathogenic and non-pathogenic human traits. L2G prioritization highlights traits linked to unique independent signals identified as haplotypes associated with polyQ disorder genes extending beyond the primary traits directly caused by the pathogenic CAG repeat expansions in these the genes.

| Signal (Tag variant)<sup>a</sup> | Reported trait (PMID) | Variant ID | Annotation<sup>b</sup> | Effect AF EUR<sup>c</sup> | GWAS index *P*-value | GWAS index effect (95% CI) | L2G | CADD score |
| --- | --- | --- | --- | --- | --- | --- | --- | --- |
| *ATXN1*: haplotype one (rs719316) | Multiple sclerosis (31604244) | rs719316 | intronic | T: 0.529 | 2.00x10<sup>-13</sup> | 1.07 (1.05-1.09)<sup>d</sup> | 0.79 | 5.92 |
| *ATXN1*: haplotype two (rs17603856) | Systemic lupus erythematosus (27399966) | rs17603856 | intronic | G: 0.346 | 3.00x10<sup>-12</sup> | 0.88 (0.85-0.91)<sup>d</sup> | 0.79 | 10.94 |
| *ATXN1*: haplotype three (rs909788) | Cognitive ability, years of educational attainment or schizophrenia (31374203) | rs6459480 | intronic | A: 0.509 | 2.00x10<sup>-10</sup> | NA | 0.69 | 1.82 |
|  | Household income (31844048) | rs6459480 | intronic | A: 0.509 | 5.00x10<sup>-10</sup> | -0.01 (-0.02--0.01) | 0.70 | 1.82 |
|  | Educational attainment (30038396) | rs9297016 | intronic | G: 0.576 | 1.00x10<sup>-09</sup> | -0.01 (-0.01--0.006) | 0.68 | 2.57 |
|  | Educational attainment: years of education (27225129) | rs7772172 | intronic | G: 0.576 | 1.00x10<sup>-08</sup> | -0.01 (-0.02--0.01) | 0.68 | 5.02 |
|  | Educational attainment<br>(years of education)<br>(30038396) | rs909788 | intronic | C: 0.545 | 4.00x10<sup>-08</sup> | -0.01 (-<br>0.011--<br>0.005) | 0.77 | 5.75 |
| *ATXN1*: haplotype<br>four (rs6459472) | Central corneal thickness<br>(32528159) | rs6459472 | intronic | G: 0.573 | 3.00x10<sup>-09</sup> | NA | 0.72 | 2.97 |
| *ATXN1*: haplotype<br>five (rs9367926) | Male-pattern baldness<br>(30573740) | rs6915310 | intronic | T: 0.182 | 5.00x10<sup>-20</sup> | 0.04 (0.03 -<br>0.05) | 0.72 | 4.27 |
|  | Balding type 1<br>(30595370) | rs9367926 | intronic | G: 0.180 | 6.00x10<sup>-12</sup> | NA | 0.71 | 6.59 |
| *ATXN1*: haplotype<br>six (rs2237199) | Subcutaneous adipose<br>tissue attenuation<br>(27918534) | rs2237199 | intronic | A: 0.112 | 1.00x10<sup>-08</sup> | 5.67 (3.73 -<br>7.61) | 0.57 | 3.49 |
| *ATXN1*: haplotype<br>seven (rs7770062) | Atrial fibrillation<br>(30061737) | rs73366713 | intronic | A: 0.131 | 1.53x10<sup>-25</sup> | 0.90 (0.88-0.92)<sup>d</sup> | 0.55 | 2.84 |
|  | Atrial fibrillation<br>(29892015) | rs7770062 | intronic | A: 0.131 | 9.00x10<sup>-21</sup> | 0.90 (0.88-0.92)<sup>d</sup> | 0.54 | 2.33 |
| *ATXN1*: haplotype<br>eight (rs3819405) | Breast cancer (29059683) | rs3819405 | intronic | T: 0.340 | 1.65x10<sup>-08</sup> | 0.96 (0.95-0.97)<sup>d</sup> | 0.53 | 12.10 |
| *ATXN2*: haplotype<br>one (rs653178) | Celiac disease (20190752) | rs653178 | intronic | T: 0.534 | 6.00x10<sup>-14</sup> | 0.83 (0.78-0.87)<sup>d</sup> | 0.51 | 1.59 |
| *ATXN2*: haplotype<br>two (rs848130) | Type 1 diabetes<br>(34012112) | rs848130 | intronic | A: 0.192 | 1.60x10<sup>-14</sup> | 0.85 (0.82-0.89)<sup>d</sup> | 0.67 | 2.02 |
| *ATXN7*: haplotype<br>one (rs832190) | Schizophrenia (31740837) | rs832190 | intronic | T: 0.603 | 4.00x10<sup>-08</sup> | NA | 0.51 | 0.58 |
| *ATXN7*: haplotype two (rs3821902) | Breast cancer (29059683) | rs3821902 | intronic | G: 0.150 | 2.99x10<sup>-12</sup> | 1.06 (1.05-1.08)<sup>d</sup> | 0.54 | 0.81 |
| *ATXN7*: haplotype three (rs13434089) | Type 2 diabetes (32541925) | rs13434089 | intronic | C: 0.141 | 4.00x10<sup>-31</sup> | -0.06 (-0.07--0.05) | 0.55 | 4.59 |
| *CACNA1A*: haplotype one (rs5021328) | Appendicular lean mass (33097823) | rs5021328 | intronic | C: 0.644 | 6.60x10<sup>-09</sup> | -0.01 (-0.02--0.01) | 0.86 | 0.63 |
| *CACNA1A*: haplotype two (rs16003) | Positive affect (30643256) | rs16003 | intronic | T: 0.463 | 3.00x10<sup>-08</sup> | 0.01 (0.00 - 0.01) | 0.84 | 8.26 |
|  | Depressive symptoms (30643256) | rs16003 | intronic | T: 0.463 | 3.00x10<sup>-08</sup> | 0.01 (0.00 - 0.01) | 0.84 | 8.26 |
| *HTT*: haplotype one (rs363096) | Noncognitive aspects of educational attainment (33414549) | rs363096 | splice | C: 0.591 | 1.00x10<sup>-12</sup> | 0.05 (0.04 - 0.06) | 0.72 | 6.29 |
|  | Educational attainment: years of education (30595370) | rs363096 | splice | C: 0.591 | 1.00x10<sup>-10</sup> | NA | 0.74 | 6.29 |
| *HTT*: haplotype two (rs61348208) | Depression (30718901) | rs7685686 | intronic | G: 0.433 | 6.00x10<sup>-15</sup> | 0.983 (0.979-0.987)<sup>d</sup> | 0.59 | 6.39 |
|  | Frailty index (34431594) | rs82334 | intronic | C: 0.323 | 3.10x10<sup>-10</sup> | -0.02 (-0.03--0.02) | 0.54 | 4.13 |
|  | Parental lifespan (30642433) | rs61348208 | intronic | T: 0.389 | 6.00x10<sup>-09</sup> | 0.23 (0.15 - 0.31) | 0.63 | 5.46 |
|  | Gastroesophageal reflux disease (34187846) | rs7685686 | intronic | G: 0.433 | 1.10x10<sup>-08</sup> | 0.97 (0.96-0.98)<sup>d</sup> | 0.54 | 6.39 |
|  | Aging traits: health span, parental lifespan or longevity (32678081) | rs61348208 | intronic | T: 0.389 | 3.00x10<sup>-08</sup> | NA | 0.62 | 5.46 |
|  | Vertex-wise sulcal depth (34910505) | rs2071703 | intronic | C: 0.325 | 3.00x10<sup>-08</sup> | 5.57 (3.60 - 7.54) | 0.6 | 2.52 |
| **HTT**: haplotype three (rs113928896) | Highest math class taken (30038396) | rs113928896 | intronic | T: 0.143 | 2.00x10<sup>-09</sup> | 0.02 (0.01 - 0.02) | 0.55 | 0.32 |
| **HTT**: haplotype four (rs362307) | Automobile speeding propensity (30643258) | rs362307 | UTR | T: 0.063 | 6.00x10<sup>-14</sup> | -0.03 (-0.04--0.02) | 0.51 | 3.23 |
|  | Worry/vulnerability: special factor of neuroticism (30867560) | rs362307 | UTR | T: 0.063 | 1.00x10<sup>-09</sup> | 0.01 (0.005 - 0.011) | 0.54 | 3.23 |
|  | Walking pace (33128006) | rs362307 | UTR | T: 0.063 | 1.00x10<sup>-09</sup> | -0.01 (-0.02--0.01) | 0.54 | 3.23 |
|  | Type 2 diabetes (30297969) | rs362307 | UTR | T: 0.063 | 1.00x10<sup>-09</sup> | 1.08 (1.05-1.11)<sup>d</sup> | 0.54 | 3.23 |
|  | Predicted visceral adipose tissue (31501611) | rs362307 | UTR | T: 0.063 | 2.00x10<sup>-09</sup> | 0.03 (0.02 - 0.04) | 0.54 | 3.23 |
| **HTT**: haplotype five (rs6828882) | Monobrow (27182965) | rs6828882 | intronic | G: 0.101 | 1.00x10<sup>-22</sup> | NA | 0.53 | 8.54 |
| **HTT**: haplotype six<br>(rs2798297) | Metabolic biomarkers<br>(33980691) | rs2798297 | intronic | A: 0.353 | 1.00x10<sup>-24</sup> | NA | 0.52 | 2.08 |

Haplotype, Group of variants in high LD with one another. Tag variant, Variant proxy for all variants in high LD at a haplotype. <sup>a</sup>LDlink SNPclip pruned signals using an r2 of 0.5 in the 1000 Genomes Project European superpopulations. <sup>b</sup>Ensembl VEP ‘most severe’ annotations. <sup>c</sup>1000 Genomes Project European superpopulation effect allele frequency. <sup>d</sup>Odds ratio (all other effects are beta estimates). **Abbreviations:** AF, Allele frequency; CADD, Combined Annotation Dependent Depletion; EUR, 1000 Genomes European super-population; L2G, Open Targets locus-to-gene scoring metric; NA, data Not available; PMID, PubMed Identifier; UTR, Untranslated region.

### Profiling polyQ disorder genes for severe Mendelian diseases beyond canonical polyQ disorders

A subset of the polyQ disorder genes (i.e., *AR, ATN1, ATXN2, CACNA1A, HTT, TBP*) had phenotype entries in the OMIM database beyond the primary polyQ disorders (**Table 2**). Rare Mendelian disorder associations included androgen insensitivity and hypospadias related to *AR*, and neurological conditions linked to *HTT* (i.e., neurodevelopmental disorder: Lopes-Maciel-Rodan syndrome), *CACNA1A* (i.e., developmental and epileptic encephalopathy) and *ATN1* (i.e., congenital hypotonia, epilepsy, developmental delay, and digital anomalies). *ATXN2* and *TBP* were also associated with susceptibility to Parkinson’s disease, a common neurological disorder, while *ATXN2* was also associated with susceptibility to amyotrophic lateral sclerosis (ALS), a neurodegenerative disease. Finally, *AR* was also associated with prostate cancer susceptibility in OMIM. These findings illustrate that therapeutic polyQ disorder gene knockdown may impair critical developmental or neurological pathways.

**Table 2.**
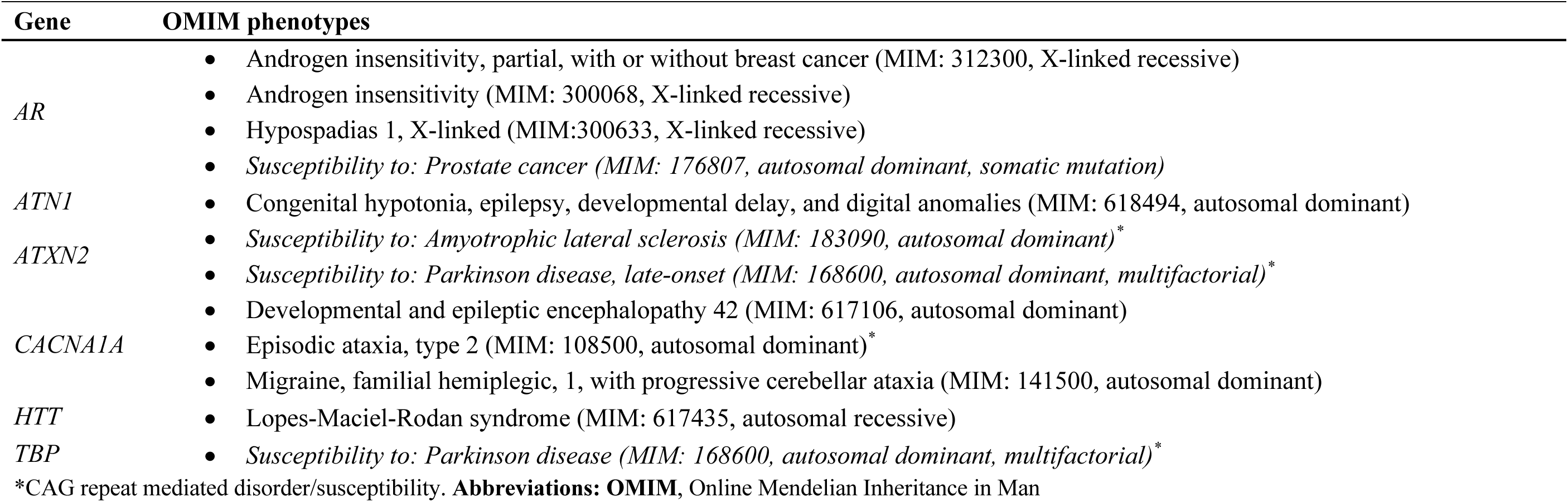
Non-canonical disease associations of polyQ disorder genes potentially informs off target effects of therapeutic knockdown of pathogenic disease genes. Six polyQ disorder genes have OMIM phenotype entries outside of their main polyQ disorder. These phenotypes range from endocrine and metabolic conditions (e.g., androgen insensitivity), to neurological and neurodevelopmental disorders (e.g., developmental and epileptic encephalopathy, Lopes-Maciel-Rodan syndrome), and susceptibility to complex diseases.

### Analysis of human genomic safety profiles and the theoretical druggability of polyQ disorder genes highlights potential safety concerns associated with therapeutic targeting

We evaluated genomic features that predict clinical trial success to better understand the theoretical implications of therapeutically perturbing polyQ disorder genes. Most of these genes showed unfavorable profiles based on multiple criteria (**Figure 3**) using thresholds defined by the original study that identified associations with these features and clinical trial stoppage.^35^ Notably, four genes (i.e., *ATN1, ATXN1*, *ATXN7* and *HTT*) ranked unfavorably for the three categories associated with clinical trial stoppage due to safety concerns (i.e., genetic constraint, broad expression, and several interactors). Further, all polyQ disorder genes displayed at least one unfavorable signal for one of these three features. All polyQ disorder genes, except for *THAP11*, displayed several molecular interactors (i.e., more than ten protein-protein interactions in the IntAct database). This was especially apparent for *HTT*, *ATXN1*, *ATN1* and *ATXN3*, which showed over 100 high-confidence interactions (i.e., ten times above the threshold used for classification). While five polyQ disorder genes fell within the *‘druggable any’* category, this metric indicates whether the gene can be theoretically perturbed, not an indicator of safety. Compared to HD age-of-onset modifier genes, polyQ disorder genes displayed significantly more unfavorable features associated with an increased safety risk (*P*=7.03×10^-3^; mean HD modifiers=1.0, *n*=9; mean polyQ disorders=2.2, *n*=10; effect size *r*=0.628, large).

**Figure 3.**
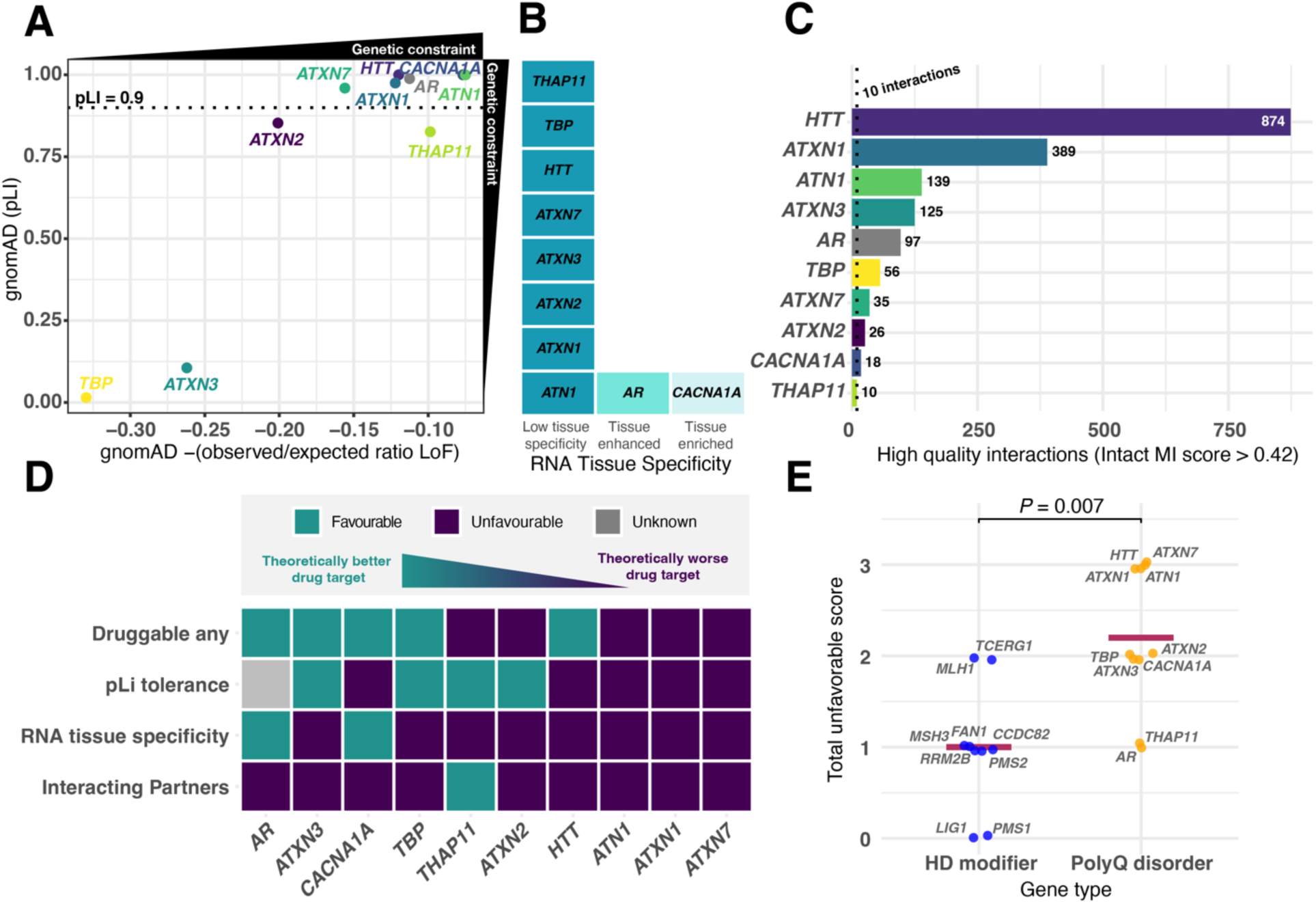
Genetic features associated with clinical trial success suggest that polyQ disorder genes may not be favorable therapeutic targets. (**A)** Genetic constraint features, gnomAD probability of loss-of-function intolerance (pLI) and negative observed vs expected loss-of-function variant ratio, indicate high levels of the constraint of the polyQ disorder genes. pLI allows non-constrained genes (pLI<0.1) to be differentiated from highly constrained ones (pLI>0.9). The observed/expected ratio is a quantitative measure of how tolerant the genes are to certain classes of variations. (**B)** Tissue specificity of the polyQ disorder genes, with *AR* and *CACNA1A* displaying tissue-enhanced and tissue-enriched, respectively, while others show low specificity. (**C)** PolyQ disorder genes interact with several other proteins and only *THAP11* displays less than 11 interacting partners (associated with decreased levels of clinical trial stoppage). (**D)** Aggregated summary of human genomic features used to rank the theoretical druggability and genomic safety of polyQ disorder genes. Genes ranked more unfavorable related to theoretical druggability and genomic safety, such as *HTT*, *ATN1*, *ATXN1* and *ATXN7*, are on the right of the plot. Individual favorable metrics are represented by the teal green tiles, while the deep purple tiles represent unfavorable features. Favorable/unfavorable classification thresholds are based on metrics identified by original study where features associated with clinical trial stoppage were identified (Razuvayevskaya *et al.* 2024 *Nat Gen*). **(E)** PolyQ disorder genes (yellow) display significantly more (*P*=0.007) unfavorable genomic safety scores based on constraint, tissue specificity and interactions (maximum score=3, increased risk of clinical trial stoppage) than Huntington disease modifier genes (blue). Mean scores are indicated by red line (mean polyQ=2.2, mean HD modifier genes=1.0).

## Discussion

Gene knockdown strategies targeting the primary disease-causing genes in polyQ disorders are being extensively explored as potential therapies in these repeat expansion disorders.^3,5,6,36^ However, these strategies have been unsuccessful in polyQ clinical trials, failing to achieve the desired therapeutic value and resulting in adverse safety profiles. To better understand these challenges, we systematically profiled polyQ disorder genes using large-scale genomic databases, highlighting their pleiotropic roles and identifying genomic features associated with increased clinical trial failure. We demonstrated that polyQ disorder genes might be unfavorable targets since they are associated with genomic features linked to an increased failure of drug targets in clinical trials. Notably, *ATN1* (DRPLA), *ATXN1* (SCA1), *ATXN7* (SCA7), and *HTT* (HD) had particularly deleterious risk profiles in our analyses.

Our analysis of human GWAS revealed that polyQ disorder genes display extensive pleiotropy beyond their canonical roles in polyQ-related neurodegeneration, indicating potential on-target effects of therapeutic knockdown.^37^ For example, *HTT, ATXN1, ATXN7* and *CACNA1A* were associated with neuropsychiatric disorders such as depression and schizophrenia. Additionally, metabolic traits like type 1 and type 2 diabetes were linked to *ATXN2* and *ATXN7*, which is in line with the fact that metabolic dysfunction is observed in several polyQ disorders.^38^ *ATXN7* was exclusively associated with pathogenic traits, including schizophrenia, diabetes, breast cancer, and cataracts, highlighting the potential risks of targeting this gene. Our OMIM analyses also revealed disease links to polyQ disorder genes outside canonical repeat-mediated disorders. For example, *ATN1* and *HTT* have been linked to neurodevelopmental disorders, while *CACNA1A* has been linked to channelopathies with diverse neurological manifestations. However, susceptibility to Parkinson’s disease and ALS are findings correlated with longer CAG repeats,^39^ suggesting possible benefits of mutation-specific knockdown related to these phenotypes.

PolyQ disorder genes were also linked to critical non-pathogenic phenotypes through GWAS. We have previously reported that common *HTT* genetic variation is associated with important neurobiological traits such as educational attainment and longevity,^15^ which we replicate here. This study showed that *ATXN1* is also linked to similar traits, including educational attainment and cognitive abilities. When comparing the variant effects between related GWAS and GTEx expression, alleles linked to lower *ATXN1* expression were also associated with lower educational attainment and decreased cognitive abilities. These findings suggest that therapeutically targeting the wild-type versions of these genes might inadvertently affect cognitive function, which should be assessed during future clinical trials. However, a key limitation of this investigation is that we cannot currently infer the direction of effect for all polyQ gene-GWAS trait pairs. Future studies should systematically address this gap using high-throughput approaches (e.g., CRISPR screens and functional assays).

In addition to individual gene-trait associations, network analyses revealed that specific trait associations were shared between polyQ disorder genes, with *ATXN1*, *ATXN2,* and *HTT* displaying the greatest number of shared interactions. For example, associations with metabolic traits (type 2 diabetes and cholesterol levels) were shared by *HTT, ATXN2* and *ATXN7* and susceptibility to breast cancer was shared by *ATXN1* and *ATXN7.* These analyses indicate that specific polyQ disorder genes may be involved in common biological pathways, as previously reported by others.^40,41^

While half of the polyQ disorder genes were found to be potentially challenging to target therapeutically with traditional approaches, RNA-targeted therapeutics provide a flexible solution by enabling the targeting of any gene product. Nonetheless, our human genomic safety profile analyses revealed that polyQ disorder genes consistently rank unfavorably for predictors of clinical trial success,^19^ which could provide some insight into the mechanisms driving gene-lowering trial failures.^6,7,42^ Specifically, *ATN1*, *ATXN1*, *ATXN7* and *HTT* ranked poorly across all safety-related categories. These four genes are integral to transcriptional regulation and are, therefore, essential for several critical physiological processes.^38^ Therapeutic targeting of these genes, especially using approaches that do not specifically target mutant versions of the related gene products, could therefore disrupt several important processes, leading to adverse effects.

Most of the polyQ disorder genes (i.e., 80%) displayed broad expression patterns, with this profile of expression associated with increased trial stoppage risks [reported odds ratio (OR): 1.29 95%; confidence interval (CI): 1.19-1.39].^19^ Additionally, half of the polyQ disorder genes are highly constrained, with LoF intolerance (pLI>0.9) also associated with an increased risk of trial stoppage (reported OR: 1.37; 95% CI: 1.27-1.48).^19^ Further, all polyQ disorder genes, except for *THAP11*, displayed more than ten high-confidence protein-protein interactions, indicating their central physiological roles – an effect that can, in part, be attributed to the polyQ repeats themselves.^43^ Specifically, genes with between 11 and 20 partners (e.g., *CACNA1A*) are at an increased risk of clinical trial stoppage (reported OR: 1.31; 95% CI: 1.19-1.44),^19^ while those with >20 partners (i.e., 80% of polyQ disorder genes) are at an even greater risk (reported OR: 1.38; 95% CI: 1.26-1.52).^19^

Finally, when we compared the genomic safety predictions for polyQ disorder genes to those of genetic modifiers of HD onset age, we demonstrated that the modifier genes showed more favourable genomic safety scores. This provides additional support for prioritizing these modifiers in future therapeutic research, especially in HD. Beyond their favourable drug target profiles, a benefit of modifier-based therapeutic targeting is the potential for cross-repeat expansion disorder relevance due to shared pathobiological mechanisms of these modifier genes (e.g., somatic repeat instability).^34,44–47^ This may also be useful for rarer repeat expansion disorders, where assembling large cohorts of individuals required for a disorder-specific clinical trial may be difficult.^36^ While these modifier genes were identified by GWAS-based approaches, which typically yield associations with modest to small effect sizes with traits being studied,^34^ the effect of therapeutically targeting these genes may have a larger impact on disease course. This is illustrated by the fact that several GWAS have confirmed the involvement of variants in known drug targets for numerous diseases.^48^ For example, *DRD2*, the target for antipsychotics, has now been robustly associated with schizophrenia risk through GWAS, although with a small effect (rs2514218: GWAS odds ratio = 1.08)^49^.

In conclusion, our findings indicate that the pleiotropic roles and adverse genomic profiles of polyQ disorder genes might diminish their potential as therapeutic targets for their respective conditions. However, it should be noted that these genes do have human genetic evidence for involvement in their primary diseases, a feature associated with increased clinical trial success.^13^ Therefore, polyQ disorder genes should not be entirely abandoned as therapeutic targets, especially with some encouraging results from recent early clinical trials.^50^ Instead, increased focus should be placed on assessing age-dependent implications of knockdown, as well as therapeutic approaches aimed at mutant allele-specific perturbation and gene editing. In parallel, building on our findings that genetic modifiers of the age of HD onset have favourable genomic safety scores, future research should intensively investigate therapeutics that target repeat expansion disorder genetic modifiers. In line with this, the promising role of therapeutic targets that prevent somatic expansions has been further emphasized by recent reports that show that the impact of the expanded repeat only becomes evident late in the lifetime of vulnerable cell types.^51^ Taken together, this study has revealed the importance of incorporating diverse human genomics data into drug development pipelines to develop safer and more effective treatments.

## Acknowledgements

We thank the Natural Sciences and Engineering Research Council of Canada through the Canada Research Chairs Initiative and Discovery Grant Program, as well as the Health Sciences Centre Foundation Winnipeg, for the financial support provided to GEBW. BID was also supported through the Canadian Institutes of Health Research Canada Research Chairs Initiative. The Research Manitoba Graduate Studentship and Rady Faculty of Health Sciences Student Top-Up supported NKL.

## Author contributions

NKL performed the formal analyses and wrote the manuscript. GEBW and BD designed the study and wrote and edited the manuscript. All authors agreed on the final version of the paper.

